# Fall Injury Avoidance Strategy Scale (FIAS) – Development and Validation of a Scale to Quantify Fall-Related Protective Movements

**DOI:** 10.1101/2025.07.22.25331998

**Authors:** Lingjun Chen, Tobia Zanotto, Joo Hyun Lee, James Fang, Abbas Tabatabaei, Neil Alexandar, Jacob Sosnoff

**Author notes:** Corresponding author: Jacob J. Sosnoff, PhD Department of Physical therapy, Rehabilitation Science, and Athletic Training, School of Health Profession, University of Kansas Medical Center, 3901 Rainbow Blvd, Kansas City, KS, 77170, United States.

## Abstract

**Objectives:** To develop and validate the Fall Injury Avoidance Scale (FIAS), a novel video-screening tool to assess protective movements and injury risk during standing-height falls among older adults.

**Design:** Secondary analysis of video-recorded experimental falls.

**Setting:** Laboratory setting.

**Participant:** Twenty-three older adults (aged 66–84 years; 4 males, 19 females) at risk of injurious falls.

**Intervention:** Not applicable.

**Main Outcome measures:** Participants experienced a total of six standardized experimentally-induced (backwards, left, and right directions). Fall movements were recorded using a motion capture system and commercially-available cameras. Hip and head peak acceleration were extracted from custom MatLab scripts. FIAS score (sum of 4 evidence-based protective fall movements, range 0–8) and the occurrence of head impact were determined by two independent raters using standardized and structured questionnaire. Inter-rater reliability, construct validity, responsiveness, and diagnostic utility of FIAS score were evaluated.

**Results:** A total of 274 experimental falls were analyzed. FIAS score indicated excellent inter-rater reliability (ICC(2,k) = 0.85 (95%CI [0.81–0.88], p<0.001). The higher FIAS scores were significantly associated with the lower hip peak acceleration (B=-1.01, 95%CI [-1.47, -0.56], p<0.001) and head peak acceleration (B=-6.08, 95%CI [-709, -5.06], p<0.001), and lower odds of head impact (Odds Ratio [OR]=0.20, 95%CI [0.12, 0.33], p<0.001). FIAS score also demonstrated significant responsiveness to the training-induced changes (F=18.74, p<0.001). Diagnostic analysis suggested that FIAS score ≥ 6 showed the high specificity (95.5%) for ruling out head impact falls.

**Conclusions:** FIAS is a reliable, valid, and responsive tool to assess the protective fall movements and injury risk among older adults, which has potential utility in screening and prevention of fall-related injuries.

## Introduction

Falls are the leading cause of injuries leading to emergency department visit, hospitalization, and injury-related death among older adults in the United States (1). ∼30% of community-dwelling older adults aged 65+ falling annually (2). The US Preventive Services Task Force (USPSTF) concluded exercise and multifactorial interventions only provide small to moderate net benefit in reducing fall-related injuries and morbidity in older adults (1), underscoring the need for novel strategies.

Injuries primarily result from the fall impact force exceeding tissue strength (3–9). Protective movements during fall descent such as squatting (8, 10), rotation/rolling (7, 11), upper limb movements (6, 12, 13), and chin tuck (8), can reduce the impact forces to vulnerable areas during experimental falls. Teaching these evidence-based protective movement strategies can reduce both impact forces and the occurrence of head impact among older populations (6–9). The performance of protective fall movement may be indicative of fall-related impact severity and injury risk.

However, assessing fall movement strategies typically involves the complex and time-consuming biomechanical analyses, and costly laboratory-based motion capture system (6–9), limiting clinical implementation. A more accessible, ‘low-tech’ approach is needed. Video analysis using commercially available cameras offers a feasible and practical solution in real-world settings. Standardized video analysis can reliably evaluate movements of key fall phases – onset, descent, and impact (8, 14–16), and can extract kinematic outcomes comparable to traditional biomechanical analysis (17, 18). Video analysis has been used to investigate the clinical and situational factors related to falls and associated injuries (19–22). It has yielded valuable insights into specific protective movement strategies, such as body rotation (23, 24), and upper limb movements (25, 26).

Existing video-based tools primarily relied on the binary categorization of single movement strategies (presence/absence) (23–26), missing the combined effect of multiple protective movement strategies. To date, no tool has encompassed multiple protective movement strategies for comprehensive evaluation of fall-related impact severity and injury risk, a practical and inclusive approach is needed.

This study aimed to develop and validate a clinically feasible video-analysis tool, the Fall Injury Avoidance Strategy Scale (FIAS), to assess protective movements and injury risk during standing-height falls. FIAS was designed to assess the quality of four evidence-based protective movements: squatting, rotation/rolling, protective upper limb movement, and chin tuck (5, 8, 10, 11, 23, 26). We hypothesize that FIAS would be a reliable and valid, with higher FIAS scores associated with lower head/hip kinematics and reduced odds of head impact (concurrent criterion validity), sensitive to the protective movement training (known-group validity), and unique contributions of each movement strategies (content validity and structural integrity).

## Methods

### Study Design, Participants, and Falling Safely Training (FAST) Program

This study is a secondary analysis of kinematics and video recordings of experimentally-induced falls from a randomized control trial (NCT05260034, R21AG073892). Study procedures were approved by the Institute Review board at the University of Kansas Medical Center (STUDY00147362), and written consent was obtained from all participants.

Eligibility criteria have been previous published (27). In brief, community-dwelling older adults aged 65 and over were included if they met these key inclusion exclusion criteria: 1) at risk of injurious falls, as indicated by meeting at least one of the following self-reported criteria (28): 1a) history of fall-related injury in the past 12 months; 1b) history of 2 or more falls in the past 12 months; or 1c) concerns of falls; 2) balance impairment, as indicated by the performance of <10 seconds on the unipedal stance test (29). Participants were excluded if they met any of key exclusion criteria: 1) history of fragility fracture; 2) bleeding risk or currently taking anticoagulants; 3) previous training experience with fall techniques (e.g., tumbling, gymnastics, or martial arts).

Eligible participants were randomly assigned to either the intervention group (FAST) or the control group (modified Otago Exercise Program, OEP). Both groups completed 4-weeks training (eight 30-minutes sessions in total) with experienced trainers (detailed training protocol published previously) (27). Participants in the FAST group received training in protective fall movements, while OEP group received traditional balance training. Falls recorded during baseline and post-training visits from 23 participants were included in the analysis (FAST=11; OEP=12), including 137 baseline falls (FAST=65, OEP=72) and 137 acquisition falls (FAST=65, OEP=72).

### Experimentally Induced Falls

During each visit, participants completed six standardized experimental falls: two falls in each directions (backward, left, and right) (27), with the order randomized across participants.

Participants wore protective gear including hip protectors, wrist guards, and a lightweighted helmet. The experimental setup included a body weight support system, an inextensible tether, a mechanical catch, a harness, and a padded landing system (as described in the previous publication) (27). To induce the experimental fall, participants were gradually leaned over 10° from the vertical by adjusting the inextensible tether until fully supported by the weight support system.

At the target leaning angle, participants were instructed to cross their arms with eyes closed to standardize the initial fall movements and minimize the prediction of the exact fall onset. Participants were provided standardized instruction to ‘landing on the mat in the way that feels the most comfortable and natural to you’. The mechanical catch was then released at pseudorandom intervals (10 to 30 seconds) without a countdown to simulate unexpected falls (Figre 1).

### Kinematics Measures of Experimental Falls

Each experimental fall was recorded by an eight-camera motion capture system (Cortex, MotionAnalysis Corp., USA) with 100Hz sampling rate. As described in the published protocol (27), raw kinematics data was post-processed in MatLab to extract the head peak acceleration (𝐴𝑐𝑐*_head_*) and hip peak acceleration (𝐴𝑐𝑐*_hip_*). Of the 274 total falls, 7 falls were excluded from kinematics-related statistical analyses due to key marker loss (Baseline: 6 falls [FAST: 3, OEP: 3]); Acquisition: 1 fall [OEP]).

### Video Recording and Analysis of Experimental Falls

Video recordings of experimental falls were obtained using 2 iPads (Apple Inc., Cupertino, CA, USA) positioned ∼2 meters perpendicular to the experimental fall setup to best capture the descent plane. Videos had at least 1,080 * 1,920 a resolution at 30 fps. Two trained researchers independently screened individual video recorded experimental falls, with disagreements resolved by consensus. All 274 experimental falls video-recorded were screened, and included in the statistical analyses.

### Occurrence of head impact

As described previously (26), the occurrence of head impact was determined based on if the head contacted with the landing surface, AND whether that contact led to significant mat deformation and/or head rebound.

### FIAS Scoring during experimental falls

Fall Injury Avoidance Strategy Scale (FIAS) was developed to qualitatively evaluate if the protective movements were effectively performed during standing-height falls. The scale incorporated four evidence-based protective movement strategies known to mitigate fall-related injuries (5–8, 11, 23, 25, 26). Also, the scale was designed to screen against the fall videos recorded by commercially available cameras, requiring minimal training from the rater(s).

Specifically, FIAS assessed each of the following protective fall movements (detailed criteria in Supplementary 1: 1) active attempt of squatting (or equivalent movement to lower the center of mass) before hip impact (squatting); 2) rotation and rolling movement to disperse impact forces (rotation/rolling); 3) appropriate upper limb movement to avoid outstretched hand landing and/or achieve successful bracing (arm use); 4) active and sufficient chin tuck to preventing excessive head motion at impact (chin tuck).

Each movement was scored on a three-point scale (0 = ineffective; 1 = partially effective; 2 = effective). After the independent screening and consensus agreement, FIAS score for each fall was calculated by summing the score of the four strategies, ranging from 0 (the worst) to 8 (the best), to represent the overall injury risk of each fall. As such a higher score indicated the greater chance to avoid fall-related injury. All 274 falls were clearly video-recorded, rated, and included in statistical analyses.

### Statistical Analyses

Demographics were descriptively reported as mean and standard deviation or median and interquartile range (IQR). Between-group comparison of demographics were examined via Mann-Whiteney U or Person’s Chi-square tests. Inter-rater reliability of individual protective movement strategies was assessed using Cohen’s Kappa statistics, and total FIAS score reliability with intraclass correlation coefficient [ICC (2,1) for single measure and ICC (2, k) for average measures].

Concurrent validity of the FIAS score was evaluated using mixed-effect models. Linear mixed models were used to examine the association between FIAS score and 𝐴𝑐𝑐*_head_* and 𝐴𝑐𝑐*_hip_* (as indicative of injury risk), while a generalized linear mixed model (GLMM) with a binary logistic link for head impact occurrence.

To assess content validity, separate mixed-effects regression models examined the unique contribution of each strategy (categorical, with “ineffective” as reference). LMMs were performed for 𝐴𝑐𝑐*_head_* and 𝐴𝑐𝑐*_hip_*, and GLMM for head impact occurrence, reporting fixed effects of individual four protective strategies.

Responsiveness of the FIAS score to training-induced changes was examined using LMM to examine the FIAS score change over time between each group. To account for the clustering of falls within participants between groups and assessments, all mixed-effect linear models included participant as a random effect, with group and time (visit) as fixed effects.

Standardized receiver operating characteristic (ROC) analysis was applied to identify the overall discriminatory ability of FIAS score (Area under curve, AUC) and to identify the optimal cut-off score that ruling out the occurrence of head impact. As the standard ROC analysis produced non-integer cut-offs, the second-step manual analysis was conducted to determine the actional integer cut-offs (sensitivity, specificity, and Youden’s Index). All statistical analyses were performed in SPSS (version 30.0, IBM Corp., Armonk, NY, USA), with the significance level set at 0.05.

## Results

### Participant demographics

Participant demographics are summarized in Table 1.

**Table 1.** Participant demographics.

|  | All | OEP | FAST | p |
| --- | --- | --- | --- | --- |
| Num of participants<br>(% Female) | 23 (87.50%) | 12 (91.67%) | 11 (72.73%) | 0.23 |
| Age (yrs) | 72.83 ± 5.48 | 71.5 ± 5.14 | 74.27 ± 5.71 | 0.17 |
| Height (m) | 1.63 ± 0.08 | 1.62 ± 0.07 | 1.64 ± 0.09 | 0.56 |
| Weight (Kg) | 71.8 (67–81.20) | 76.45 (68.2–85.3) | 69.6 (64.9–74.6) | 0.23 |
| BMI (Kg/m<sup>2</sup>) | 28.25 ± 4.85 | 29.78 ± 5.80 | 26.58 ± 2.98 | 0.15 |

Legend. BMI: body mass index; FAST: Falling Safely Training; OEP: Otago Exercise Program.

### Inter-rater reliability

For the four individual protective movement strategies, the observed agreement and Cohen’s Kappa coefficient are presented in Table 2. Overall, the agreement ranged from 58.0% to 91.7%, with Kappa coefficient indicating fair to outstanding agreement across individual strategy items.

**Table 2.** Inter-rater reliability of four protective movements between two raters.

|  | % Agreement | Cohen's Kappa (95%CI) | p-value |
| --- | --- | --- | --- |
| Q1. Squatting | 77.2% | 0.51 (0.40, 0.61) | < 0.001 |
| Q2. Rolling/Rotation | 58.0% | 0.32 (0.23, 0.41) | < 0.001 |
| Q3. Arm use | 91.7% | 0.83 (0.76, 0.90) | < 0.001 |
| Q4. Chin tuck | 75% | 0.60 (0.52, 0.68) | < 0.001 |

For the FIAS score, ICC(2,1) for a single measure was 0.74 (95%CI [0.68–0.79], p<0.001), while the ICC(2, k) for the average of two measures was 0.85 (95%CI [0.81–0.88], p<0.001).

### Concurrent validity of FIAS Score

LMMs indicated the FIAS score was significantly associated with the kinematic outcomes during the experimental falls. A higher FIAS score was significantly associated with lower 𝐴𝑐𝑐*_hip_* (B=-1.01, 95%CI [-1.47, -0.56], p<0.001) and lower 𝐴𝑐𝑐*_head_* (B=-6.08, 95%CI [-7.09, -5.06], p<0.001). In addition, GLMM also revealed a significant association between FIAS score and lower odds of the head impact (Odds Ratio [OR]=0.20, 95%CI [0.12, 0.33], p<0.001).

### Content validity of FIAS

LMMs assessed the unique contribution of individual protective movement strategies to the 𝐴𝑐𝑐*_hip_* and 𝐴𝑐𝑐*_head_*. For 𝐴𝑐𝑐*_hip_*, squatting (F=4.82, p=0.009) and chin tuck (F=8.53, p<0.001) were found to be significantly associated, while rolling/rotation (F=0.22, p=0.804) and arm use (F=2.90, p=0.057) were insignificant. For squatting, compared to an ineffective strategy, both partially effective (B=–4.09, 95%CI [–7.26, –0.93], p= 0.012) and effective (B=–5.33, 95%CI [– 8.75, –1.91], p=0.002) strategies were significantly associated with lower 𝐴𝑐𝑐*_hip_*. Similarly, compared to ineffective chin tuck both effective (B=–4.39, 95%CI [–6.58, –2.19], p<0.001) and partially effective (B=–3.71, 95%CI [–5.68, –1.74], p<0.001) strategies were also associated with significantly lower 𝐴𝑐𝑐*_hip_*.

For 𝐴𝑐𝑐*_head_*, three movement strategies – rolling/ration, arm use, and chin tuck – were significantly associated with 𝐴𝑐𝑐*_head_*. For rolling/rotation (F=11.30, p<0.001), compared to an ineffective strategy, the effective strategy was associated with significantly lower 𝐴𝑐𝑐*_head_* (B=-10.06, 95%CI [-14.52, -5.60], p<0.001), while the partially effective strategy was not (B=-3.57, 95%CI [–7.40, 0.25], p=0.067). For arm use (F=18.14, p<0.001), compared to the ineffective arm movements, both partially effective (B=-8.69, 95%CI [-14.62, -2.71], p=0.005) and effective (B=-8.69, 95%CI [-12.49, -4.24], p<0.001) strategies were associated with significant lower 𝐴𝑐𝑐*_head_*. For chin tuck (F=29.73, p<0.001), compared to the ineffective chin tuck, both partially (B=-9.20, 95%CI [-13.38, -5.02], p<0.001) and effective (B=-18.26, 95%CI [- 23.02, -13.49], p<0.001) strategies were significantly associated with lower 𝐴𝑐𝑐*_head_*.

GLMM assessed the unique contribution of individual protective movement strategies to the head impact during experimental falls. Arm use (F(2, 263)=7.85, p<0.001) and chin tuck (F(2, 263)=18.86, p<0.001) were significantly associated with the lower odds of head impact, while squatting (F(2, 263)=1.03, p=0.360) and rolling/ration (F(2, 263)=1.16, p=0.36) were insignificant. For arm use, compared to an ineffective strategy, both partially effective strategy significantly reduced the odds of head impact (OR=0.03, 95%CI [0.01, 0.19], *p*<0.001). While the effective strategy yielded an extremely low estimated odds ratio (OR≈0, 95%CI [near-zero, 0.006], *p*=0.972), but the estimate was unstable with extremely wide confidence intervals, warranting cautious interpretation. For chin tuck, compared to an ineffective strategy, both partially effective (OR=0.08, 95%CI [0.002, 0.36], *p*=0.001) and effective (OR=0.001, 95%CI [<0.001, 0.006], *p*<0.001) were significantly associated with the lower odds of head impact.

### Cut-off Score for Head Impact classification

Standard ROC analysis demonstrated that FIAS score had a good discriminatory performance (Figure 1), with AUC of 0.88 (95%CI [0.84, 0.92], p<0.001). Sensitivity, specificity, and Youden’s index, accuracy for integer cut-off scores are reported in Supplementary 2. A cut-off of ≥ 5 yielded the highest Youden’s index (0.59), optimally balancing between sensitivity (76.76%) and specificity (82.0%) for head impact classification, with 78.5% overall classification accuracy.

**Figure 1.**
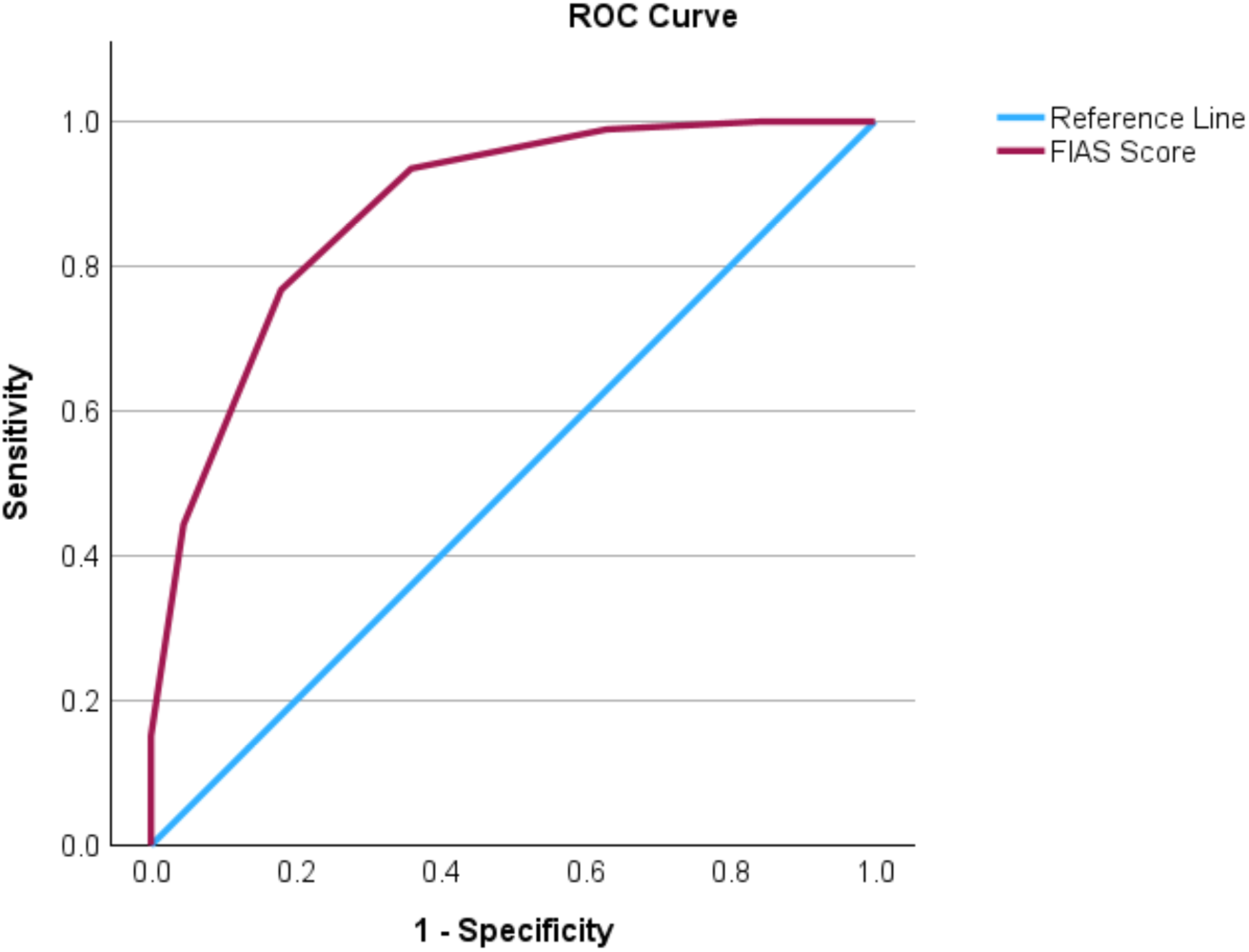
Receiver operating characteristics (ROC) curve for FIAS score in predicting head impact.

For specific clinical applications, two additional cut-offs were informative: <3 (i.e., cut-off ≥ 3) maximized sensitivity (98.9%) and yielded a high negative predictive value (NPV) for 94.3%, suggesting confidence in ruling in the risk of head impact; ≥ 6 maximized specificity (95.5%) and achieved a high positive predictive value (PPV) of 95.4%, providing strong confidence in ruling out the risk of head impact.

### Responsiveness of FIAS Score to Training

The significant interaction between time and group (F=18.74, p<0.001) suggests the change of FIAS score from baseline and post-training was significantly different between FAST and OEP groups. Specifically, the FAST group showed a significant increase in FIAS score from 3.86 (Standard Error [SE]=0.21) at baseline to 5.31 (SE=0.17) at post-training (an increase of 1.45 points); while OEP group showed a no change in FIAS score from 4.44 (SE=0.17) at baseline to 4.61 (SE=0.19) at post-training (an increase of 0.17 points) (Figure 2).

**Figure 2.**
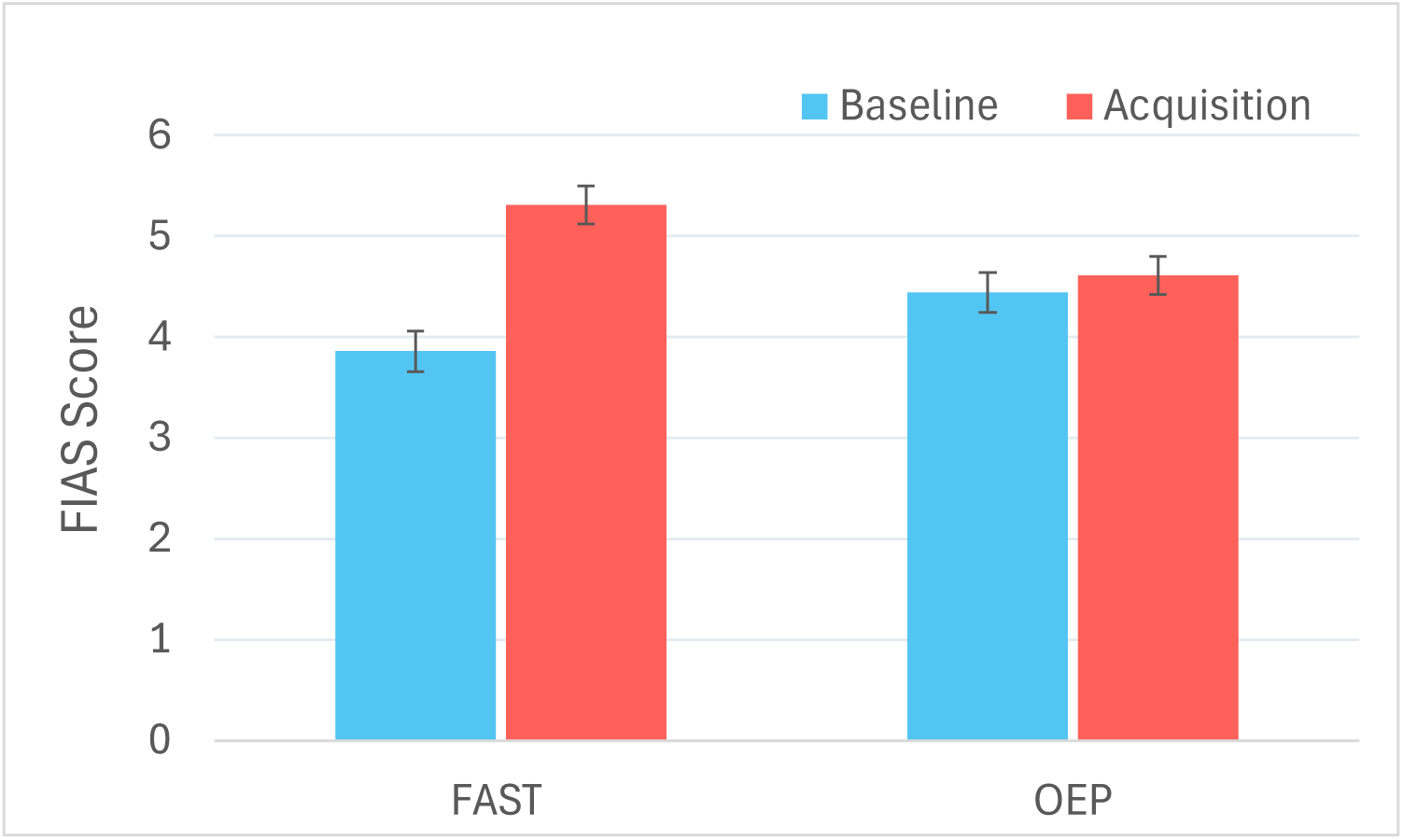
FIAS score at two groups at baseline and post-training assessment.

## Discussion

This study developed and validated the Fall Injury Avoidance Scale (FIAS), a novel video-based tool to assess protective movements and risk of fall-related injuries among older adults. FIAS demonstrated strong reliability, validity, and responsiveness, offering a practical and accessible ‘low-tech’ alternative to traditional lab-based biomechanical analysis. By integrating four evidence-based protective movements into one scoring framework, FIAS addresses a critical need for a scalable assessment tool for protective movements.

The FIAS demonstrated generally strong inter-rater reliability. Three protective movement strategies (squatting, arm use, and chin tuck) showed moderate to excellent agreement. Rolling/rotation demonstrated only fair agreement, likely as a results of the experimental paradigm which limited the opportunity for substantial rotation. Significant fall direction changes from fall onset to landing – commonly used to assess rotation in real-fall video analysis (23, 24) – were rare, likely increasing scoring variability. Despite this, the overall FIAS score remained consistent and reliable across raters, supporting the tool’s robustness.

FIAS demonstrated strong concurrent criterion validity, as evidenced by strong association between FIAS score and established biomechanical proxies for fall-related impact severity and injury risk. Specifically, a higher FIAS score showed significant association with lower 𝐴𝑐𝑐*_head_* and 𝐴𝑐𝑐*_hip_*, representing two critical body segments for impact severity (5). Higher FIAS scores were also associated with substantial reduced risk of head impact, the direct cause of head injury. Notably, each 1-point increase in FIAS score was associated with an approximately 80% reduction in head impact.

FIAS score demonstrated excellent discriminatory ability in identifying falls with or without head impact. While the cut-off score of ≥ 5 balanced sensitivity and specificity best, two additional cut-offs offered clinical relevance: <3 maximized sensitivity (98.9%) with high negative predictive value (94.3%), making it useful to identify who are at high risk of head impact (e.g., screening), while ≥ 6 maximized specificity (95.5%) with high positive predictive value (95.4%), making it useful to identify who are at low risk of head impact (e.g., evaluation of training effectiveness). Multiple thresholds enhance FIAS’s utility across diverse clinical or research purpose.

This study also provided compelling evidence for the content validity of FIAS. Previous studies indicated the protective role of various individual movement strategies (squatting, rolling/rotation, upper limb movement, and chin tuck) (6–8, 10–13, 23–26). This is the first study to comprehensively examine their distinct and collective contributions within a standardized tool. Data strongly support the inclusion of four distinct protective movements, each contributing meaningfully and independently to reducing the fall-related impact severity and injury risk.

It was found the both effective squatting and chin tuck were important to reduce 𝐴𝑐𝑐*_hip_*, as an established proxy for fall-related hip injuries. The association between squatting and hip impact severity is consistent with foundational biomechanical principles, that the active attempt of lowering center of mass before hip impact can reduce impact forces (5, 8, 10). The association between effective chin tuck and reduced 𝐴𝑐𝑐*_hip_* was unexpected. It may reflect individual demonstrating effective chin tuck adopted a generally protective strategy profile, including effective squatting during falls. This co-occurrence of multiple protective strategies warrants further investigation.

This study reinforced and expended prior findings, that rolling/rotation, upper limb movements, and chin tuck are protective for fall-related head injuries (9, 23, 26). All three movement strategies were significantly associated with the reduced 𝐴𝑐𝑐*_head_*, with a clear gradient of protection based on movement quality. Chin tuck demonstrated the strongest protective effect (>18 m/s² reduction when performed effectively). Even partially effective performance led to substantial reductions (approximately 9 m/s²). Similarly, upper limb movements and rolling/rotation yielded 3.5 - 10 m/s² reduction depending on the movement quality (the more effective the movement were, the more reduction in 𝐴𝑐𝑐*_head_*). Importantly, these biomechanical benefits translated into meaningful reduction in head impact occurrence: upper limb movements and chin tuck strategies associated with substantially lower odds of head impact (OR=0.03–0.08, partially effective; ORs<=0.001, effective). The extremely low odds estimate for fully effective strategies reflected the statistical separation in the data, where no head impacts were observed with effective upper limb movements and very few with effective chin tuck. These findings emphasize the importance of each strategy’s inclusion, support the structural integrity of FIAS, and underscore the value of teaching four protective movements to prevention fall-related injuries.

Moreover, FIAS indicated strong known-groups validity by detecting training-induced improvements, with the FAST group showing significant score increase after training. This aligns with the improved ‘mastery level’ of taught movements (9), supporting FIAS as an objective measure of objective and responsive outcome measure. Unlike existing fall competency scoring system that rely on single-rater, real-time observation of non-repeatable, self-initiated falls (often in just a second) (30), FIAS leveraged the video-recorded falls for repeatable, standardized, and multi-rater assessment for the protective movement performance.

Overall, FIAS provides a clinically meaningful, and scalable approach to assess the quality of four evidence-based protective fall movement strategies. Its structured, low-cost format makes it accessible for use in both clinical and research setting to identify older adults who lack key protective movements during falls and at high risk of fall-related injuries. While the protective training appears to be a promising approach, there remained limited data on the effect of such an intervention outside this research laboratory (9). FIAS can help to bridge the gap by supporting the implementation of screening, targeted intervention, and evaluation of future intervention on reducing fall-related injuries.

Several limitations should be acknowledged when interpreting the study findings. Firstly, the lean-and-release fall paradigm limited the reactive steps and enforced immediate descent, improving methodological consistency but reducing ecological validity (e.g., initial momentum, situational variability) (21, 22, 31). This likely restricted stepping or more pronounced rolling and rotation (13, 23), contributing to lower inter-rater reliability for that FIAS item. Future work should tailor FIAS for more ecologically valid scenarios by expanding items and refining the scoring protocols for subtle movements. Secondly, all falls were induced in a controlled laboratory setting with the simplified environmental context. Protective value of fall movements likely depend on the situational and environmental scenarios. For example, the successful bracing achieved in the current paradigm may not be as effective as holding the nearby handrails or weight-bearing objects (25). Thirdly, pooling falls in different directions (backward and sideway) may overlook direction-specific strategy effectiveness. Injury risk vary between fall direction (20, 23), and some strategies may be inherently less protective in certain directions (e.g., upper limb bracing in the backway falls) (23, 26). Forward falls were not included and warrant future investigation. Lastly, high-resolution, multi-angle video may not be feasible in a typical clinical or community setting. FIAS usability and reliability should be examined with diverse video sources, including the potential integration of AI-based movement tracking and rating to enable standardized scoring from a range of recordings (18).

## Conclusion

This study developed and validated the Fall Injury Avoidance Scale (FIAS) as a reliable, valid, and responsive tool to assess the protective fall movements and injury risk among older adults. FIAS bridges the gap between high-fidelity biomechanical analysis and scalable clinical tools. Integrating four evidence-based protective movements into one scoring system provides a novel, comprehensive approach to evaluate the quality of protective movements. FIAS has the potential to be a practical, low-cost tool to assess fall-related injury risk in various settings, which remains for further investigation.

## Funding resources

This work is supported in part by a research grant from the National Institutes of Health (1R21AG073892-01) awarded to J.J.S. J.R.F. was supported in part by the National Institutes of Health (T32HD057850). The funders have no role in study design, data collection and analysis, decision to publish, or preparation of the manuscript.

## Supporting information

Suuplementary 2

## Data Availability

All data produced in the present study are available upon reasonable request to the corresponding author

