## Supplementary 1 for "Fall Injury Avoidance Strategy Scale (FIAS) – Development and Validation of a Scale to Quantify Fall-Related Protective Movements"

**Fall Injury Avoidance Scale (FIAS)**

**Instructions:**

- Select the single most appropriate answer to each item.

**Video details**

a) Participant ID: ______________
b) Fall Direction (0 = backward, 1 = left, 2 = right) ________
c) Trial ID (1 or 2): ______________
d) Date of analysis: ______________
e) Rater: ______________

**Q1: Active attempt of squatting**

Select the answer that best describes the participant’s effort to lower the center of mass during fall descent.

0 = Ineffective: No observable attempt to flex the knees and/or hip.

1 = Partially effective: Observable flexions at knee(s) and hip but without active attempt to sit close to the heels.

2 = Effective: Observable flexions at knee(s) and hip with an active attempt to sit close to the heels.

**Q2: Rotation and/or rolling on the back motion**

Select the answer that best describes whether the participant showed the rotation and/or rolling to reduce direct vertical impact during fall.

Rotation (only in sideway falls): Torso rotation before hip impact, that the upper arm and greater trochanter not directly impact on the landing surface.

Rolling (backward and sideway falls): Rolling motion from the buttock to the upper back since hip impact.

0 = Ineffective:

- Sideway falls: No observable rotation AND rolling;
- Backward fall: No observable rolling.

1 = Partially effective:

- Sideway falls: Observable rotation OR rolling;
- Backward fall: Partial rolling (e.g., rolling stops midway before reaching the upper back).

2 = Effective:

- Sideway falls: Observable rotation AND rolling;
- Backward fall: Complete rolling from buttock to the upper back.

**Q3: Upper limb movements**

Select the answer that best describes whether the participant moved upper limb movements safely and/or effectively in bracing the fall.

0 = Ineffective:

- No observable upper limb movement (remaining in initial posture during fall), Or
- Observable upper limb movements but without contact to the landing system, Or
- Upper limbs exposed under risk of injury (landing on the outstretched hand(s)).

1 = Partially effective: Observable upper limb movement attempts and with low injury risk.

2 = Effective:

- Proper use of upper limbs in the optimal landing sequence (slapping the landing surface from upper arm, to forearm, to hand), Or
- Achieved successful bracing, evidenced by an observable space between upper torso and landing surface at fall termination.

**Q4: Sufficient head control**

Select the answer that best describes the chin tuck movement throughout the fall.

0 = Ineffective: No observable or failed chin tuck. Head significantly deviated from torso’s motion plane (approaching floor)

1 = Partially effective: Observable chin tuck. Head generally kept at/above the torso’s motion plan, but somewhat deviate towards floor.

2 = Effective: Successful chin tuck. Head kept above the torso’s motion plane throughout the fall.

**Total Score** (Sum of Q1 – Q4) = _________
