## Supplementary material for "Fall Injury Avoidance Strategy Scale (FIAS) – Development and Validation of a Scale to Quantify Fall-Related Protective Movements": Suuplementary 2

**Supplementary Table 1.** Diagnostic performance of different cut-off scores for predicting the absence of head impact

| Cut-off | Probability  (%) | Sensitivity  (%) | Specificity (%) | Youden’s Index | Accuracy (%) | PPV  (%) | NPV  (%) |
| --- | --- | --- | --- | --- | --- | --- | --- |
| $=$0 | 0.0 | 100.0 | 0.0 | 0.0 | 67.5 | 67.5 | NA |
| $\geq$1 | 3.35 | 100.0 | 5.6 | 0.06 | 69.3 | 68.8 | 100 |
| $\geq$2 | 10.92 | 100.0 | 15.7 | 0.16 | 72.1 | 71.2 | 100.0 |
| $\geq$3 | 30.27 | 98.9 | 37.1 | 0.36 | 78.8 | 76.6 | 94.3 |
| $\geq$4 | 60.58 | 93.5 | 64.0 | 0.58 | 83.9 | 84.4 | 82.6 |
| $\geq$5 | 84.48 | 76.8 | 82.0 | 0.59 | 78.5 | 89.9 | 62.9 |
| $\geq$6 | 95.07 | 44.3 | 95.5 | 0.40 | 60.9 | 95.4 | 45.2 |
| $\geq$7 | 98.56 | 15.1 | 100.0 | 0.15 | 42.7 | 100.0 | 36.2 |
| $=$8 | 99.59 | 1.6 | 100.0 | 0.02 | 33.6 | 100.0 | 32.8 |

Abbreviations: PPV: positive predictive value; NPV: Negative predictive value; NA: not applicable (due to division by zero for NPV when no true negatives are correctly predicted for the negative outcome).
